# Physical Activity Patterns Among Patients with Intracardiac Remote Monitoring Devices Before, During, and After COVID-19-related Public Health Restrictions

**DOI:** 10.1101/2021.02.27.21252558

**Authors:** Yuan Lu, Karthik Murugiah, Paul W Jones, Daisy S Massey, Shiwani Mahajan, César Caraballo, Rezwan Ahmed, Eric M Bader, Harlan M Krumholz

## Abstract

Nationwide public health restrictions due to the coronavirus disease 2019 (COVID-19) pandemic have disrupted people’s routine physical activities, yet little objective information is available on the extent to which physical activity has changed among patients with pre-existing cardiac diseases. Using remote monitoring data of 9,924 patients with pacemakers and implantable cardiac defibrillators (ICDs) living in New York City and Minneapolis/Saint Paul, we assessed physical activity patterns among these patients in 2019 and 2020 from January through October. We found marked declines in physical activity among patients with implantable cardiac devices during COVID-19-related restrictions and the reduction was consistent across age and sex subgroups. Moreover, physical activity among these vulnerable patients did not return to pre-restrictions levels several months after COVID-19 restrictions were eased. Our findings highlight the need to consider the unintended consequences of mitigation strategies and develop approaches to encourage safe physical activity during the pandemic.

## INTRODUCTION

Nationwide public health restrictions due to the coronavirus disease 2019 (COVID-19) pandemic have disrupted people’s routine physical activities, especially in urban areas.^1, 2^ Little objective information is available on the extent to which physical activity has changed among patients with pre-existing cardiac diseases, who are at high risk of adverse clinical outcomes from reduced physical activity.^3^ Pacemakers and implantable cardiac defibrillators (ICDs) implanted in cardiac patients are a reliable source of activity information through their integrated accelerometers and remote monitoring capabilities.^4, 5^ Using remote monitoring data of patients with pacemakers and ICDs living in New York City (NYC) and Minneapolis/Saint Paul (MSP), we sought to assess physical activity patterns among these patients in 2019 and 2020 from January through October.

## METHODS

A de-identified dataset of individuals aged ≥18 years from NYC and MSP with pacemakers or ICDs transmitting between January 1^st^, 2019 and October 9^th^, 2020 was derived from Boston Scientific Corporation’s LATITUDE™ database. Patient activity was measured through an accelerometer that detected the frequency and amplitude of patient motion. An algorithm specified whether the sensor exceeded a threshold of 25 milligravities, corresponding to a walking speed of 2 miles per hour, to determine a state of “active” or “not active” for a given minute. Daily activity level was defined as minutes spent in an active state per day. Sedentary status was defined as <30 minutes spent in an active state per day.

We assessed the trend in daily activity level using the 7-day moving average of activity between January 1^st^, 2020 and October 9^th^, 2020. We used data from the same months in 2019 as a comparator to account for seasonal variability and then calculated the change in activity during and after COVID-19 restrictions in 2020, overall and by age and sex. We considered 2-sided P-values <0.05 as statistically significant. Analyses were performed using R version 4.0.2. This study was approved by the Institutional Review Board at Yale University.

## RESULTS

A total of 9,924 patients from NYC (n = 6,449) and MSP (n= 3,475) were included, of whom the mean age was 73.9 [SD, 11.9] years and 42.1% were women. Before COVID-19 restrictions began in 2020, the average daily physical activity was 89.7 (± 4.7) minutes among patients in NYC and 87.5 (± 4.3) minutes among patients in MSP. Physical activity significantly declined from the week that the emergency quarantine order was issued (starting 03/07/20 in NYC and 03/13/20 in MSP) to the week that the stay-at-home restriction ended (ending 06/08/20 in NYC and 05/18/20 in MSP) in each city (P <0.001, **Figure**). Compared to data from the same period of 2019, the median reductions in physical activity were 26 minutes (27% reduction) in NYC and 15 minutes (16% reduction) in MSP, and the percentage of patients with sedentary status increased by 51.9% (27.1% vs 17.8%) in NYC and 48.5% (22.8% vs 15.3%) in MSP. In both cities, physical activity decreased the most during the first three weeks of the emergency quarantine order, and then started to slowly increase. However, five months after restrictions were lifted, activity levels had not returned to pre-pandemic levels. Compared to the same period in 2019, the 2020 daily activity level remained 13.2 minutes (84.6 vs. 97.8 minutes; p< 0.001) lower in NYC and 10.2 minutes (86.2 vs. 96.3 minutes; p<0.001) lower in MSP. This trend was noted in each of the age and sex subgroups.

**Figure.**
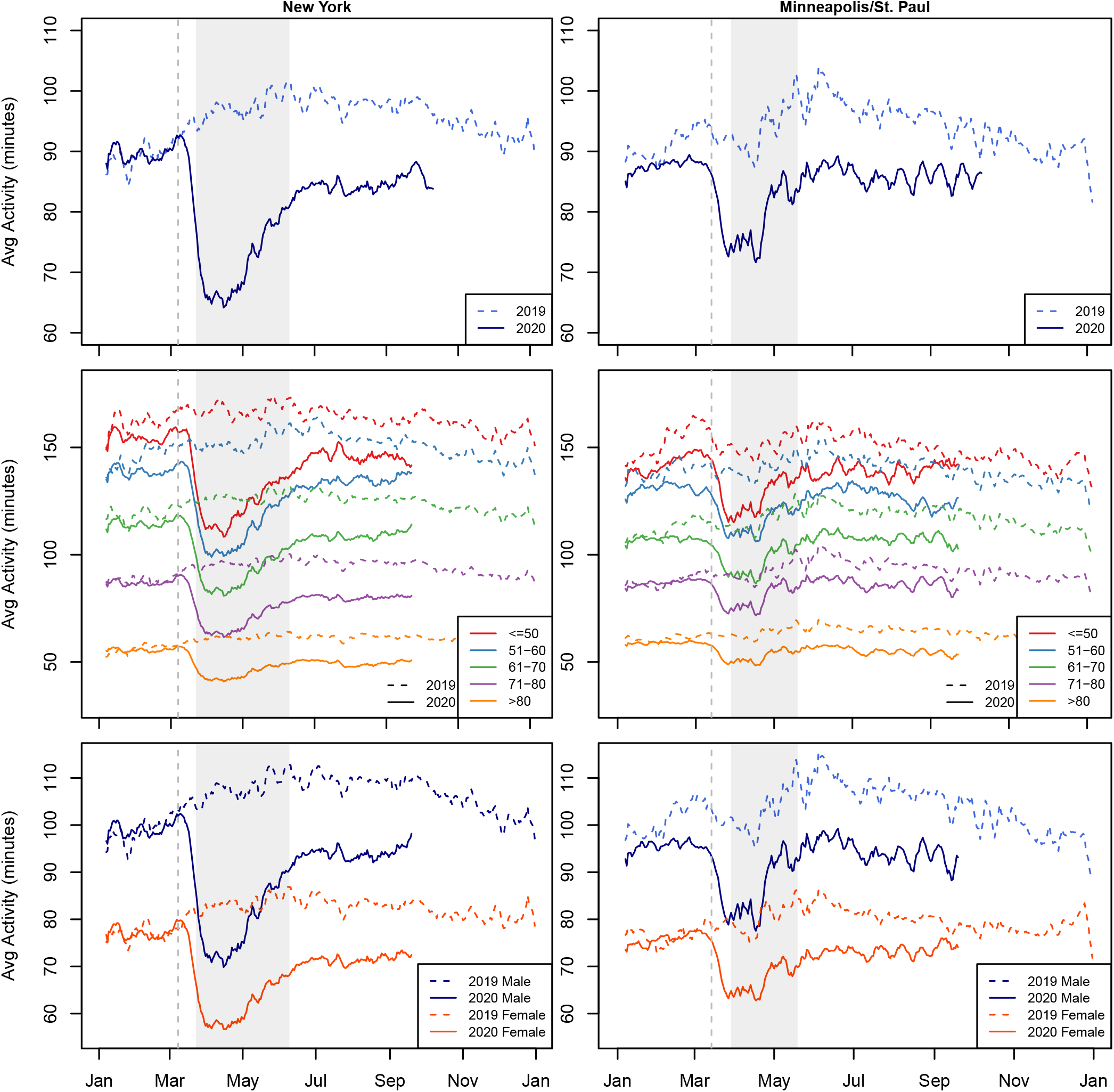
Trends in daily physical activity among patients with intracardiac remote monitoring devices in New York City and Minneapolis/Saint Paul in 2019 and 2020. The left side is New York City, and the right side is Minneapolis/Saint Paul. Top row: Overall Middle row: By Age Group Bottom row: By Gender The vertical line is the date of emergency order and the shaded area is the stay-at-home order for each city.

## DISCUSSION

We found marked declines in physical activity among patients with implantable cardiac devices during COVID-19-related restrictions and the reduction was consistent across age and sex subgroups. These findings complement reports from consumer devices that have showed rapid step-count decreases across multiple countries during the COVID-19 pandemic.^1, 2^ Moreover, physical activity among these vulnerable patients did not return to pre-restrictions levels several months after COVID-19 restrictions were eased.

This study has several implications. While the decline in physical activity during lockdown period is expected, most patients did not resume previous activity levels after the restrictions were lifted. It highlights the needs for physicians to identify patients with significant activity reductions and help them integrate simple, safe ways to stay physically active within the limits of ongoing pandemic restrictions. Potential strategies may include targeted messaging of exercise advice to these patients and promotion of at-home and outside exercise for patients used to exercising in a gym.

Limitations include the lack of information on intensity and type of activity and on other factors that may affect activity level of these patients.

In conclusion, the COVID-19 pandemic and associated public health restrictions had a significant impact on physical activity levels among patients with pacemakers and ICDs. In a pandemic that is not yet over, we need to consider the unintended consequences of mitigation strategies and develop approaches to encourage safe physical activity during these trying times.

## Data Availability

Aggregate data of this paper are available upon request to the authors.

